# A Novel Primer Probe Set for Detection of SARS-CoV-2 by Sensitive Droplet Digital PCR

**DOI:** 10.1101/2020.11.03.20224972

**Authors:** Fang Wang, Umar Pervaiz, Hongwei Tian, Algahdary Omar Ahmed Omar Mariam, Mahasin Abdallah Mohammed Hamid, Degui Wang

**Affiliations:** School of Basic Medical Sciences, Lanzhou University, Lanzhou, China; Center of Medical Experiment, School of Medicine, Lanzhou University, Lanzhou, China; Department of General Surgery, Gansu Provincial People’s Hospital

**Keywords:** Primer, Probe, SARS-CoV-2, 2019-nCov, droplet digital PCR

## Abstract

**Background:** The current increase in the spread of (SARS-CoV-2) critically needs a multitarget diagnostic assays to promote analytical sensitivity to facilitate the public health actions.

**Objective:** The aim of this study was to develop a new primer-probe set targeting N gene of SARS-CoV-2 to improve the sensitivity for detection of COVID-19(Corona Virus Disease 2019)in multiplex rRT-PCR (Reversetranscript Realtime PCR) and ddPCR (Droplet Digital PCR).

**Results:** We designed primers/probes set N(LZU3) targeting the N gene of 2019-nCov and proved its sensitivity in both rRT-PCR and ddPCR. When the quantity of template was 105 copies/reaction, the mean Ct value of N(LZU3) was 32.563, the detection rate was 91.7%. If the quantity of template was 52.5 copies/reaction, the mean Ct value of N(LZU3) was 33.835, and the detection rate was 83.3%, which were similar with that of N(CDC) and N(USA). The calculated lower limit of detection (LOD) of the new primer-probe set N(LZU3) used in rRT-PCR was 118 copies/reaction. We also did one-step ddPCR for detection the same serial dilution of RNA template. It shows good linearity for primer/probe sets N(LZU3). The calculated lower limit of detection (LOD) of N(LZU3) was 22.4 copies/reaction, which was 1.12 copies/ul.

**Conclusion:** The novel primer-probe set(LZU3) targeting N gene of SARS-CoV-2 could be both used in rRT-PCR and ddPCR with better sensitivity, furthermore, ddPCR method had higer sensitivity than rRT-PCR, hence it could significantly improve SARS-CoV-2 detection efficiency in low virus load and asymptomatic infection.

## 1. Introduction

Coronaviruses (CoVs) are one of the largest class of RNA associated viruses that are responsible for infectious disorders in human beings and mammals, the word Coronavirus derived from Latin the meaning is crown. Coronaviruses belong from the *Coronaviriade* family that has a positive-sense single-stranded RNA genome. It comprises a sizeable genome range between 26 to 32 kilobases; they have a club-shaped structure under electron microscopy. According to the distinctive genomic sequences, the *Coronavirinae* subfamily is divided into four leading subgroups: *Alpha, Beta, Gamma*, and *Delta*, coronaviruses^[1, 2]^. Previously, almost six human pathogenic coronaviruses were identified. The Novel Coronavirus SARS-CoV-2 is highly pathogenic that is caused by severe acute respiratory syndrome coronavirus 2 (COVID-19). The genome sequences have revealed that SARS-CoV-2 is closely related to SARS-CoV,it showed more than 80% similarity with the genomes of other bat derived coronaviruses^[1, 2]^.

In the interest of people’s health in January 2020, the WHO announced the pandemic of COVID-19 a Public Health Emergency of International Concern and counseled the countries with poor health systems^[3]^. According to the COVID-19 situational report 209, by 16 August 2020, coronavirus disease has spread globally in 216 countries or territories with 21 294 845 confirmed cases and 761 779 confirmed deaths^[4]^.

The COVID-19 affected individuals can be symptomatic or asymptomatic, in the symptomatic host’s clinical manifestations are visible, but there is a lack of clinical signs in asymptomatic hosts^[5, 6]^. However the early recognition of SARS-CoV-2 is difficult because its general manifestations are similar to other atypical pneumonia, the capability to verify or clear the suspected client during the worldwide epidemic scenarios is crucial especially when the clinical signs are hard to differentiate^[7]^.

At present, there are several tools and tests are suggested by health care authorities for the diagnosis of COVID-19, but previous studies mentioned that molecular diagnosis is a standard and most effective technique to examine rapidly human pathogenic viruses than the CT scan and serological approaches^[8-12]^. Moreover, coronaviruses are challenging to cultivate in cell culture and isolation of the pathogenic viruses demand safety level 3 proficiency, which is not accessible in most public health institutions, because of limitations, polymerase chain reaction (PCR) remains the most beneficial laboratory diagnostic test for COVID-19 globally^[13, 14]^.

SARS-CoV-2 genome structure was published to promote molecular diagnostic tests, which assist to develop specific sets of primers and probes to detect SARS-CoV-2^[15]^. The first assay was released on Jan 23 to target RNA-dependent RNA polymerase (RdRp), envelope (E), and nucleocapsid (N) genes of SARS-CoV-2^[14]^.

In the current situation several real-time RT-PCR and reverse transcription-PCR (RT-PCR) assays have been evaluated by using different primers and probes to examine SARS-CoV-2 with safety to decrease the possibility of false laboratory diagnosis^[14, 16, 17]^.

Although Some specific primers and probes have been formulated to target (ORF) 1ab, envelop (E) and nucleocapsid (N) gene,but the main issue in the diagnosis of COVID-19 is to increase the detection sensitivity and specificity of SARS-CoV-2^[14, 18, 19]^.

Previous reports showed that sensitivity of SARS-CoV-2 identification depends on the limit of detection (LOD) of designed primers and kit reagents. The RdRP and E genes had notable analytical sensitivity for detection whereas the N gene showed low analytical sensitivity^[13, 19, 20]^. The WHO advised to detect at least two different targets from the SARS-CoV-2 genome to diminish the possibility of false results^[19]^.

In the present study, we designed the set of primers and probes N(LZU3) targeting N gene of SARS-CoV-2 (LZU3) to developed the quantitative reverse transcriptase-polymerase chain reaction (rRT-PCR) and droplet digital polymerase reaction (ddPCR) for the detection and screening of SARS-CoV-2.

## 2. Materials and methods

### 2.1 SARS-CoV-2 In Vitro Transcribed RNA Reference Material

We used SARS-CoV-2 In Vitro Transcribed RNA Reference Material as the template in rRT-PCR and ddPCR. This material was purchased from National Institute of Metrology of China (Beijing, China). This reference material was obtained by in vitro transcription of key characteristic genes of SARS-CoV-2, including the nucleoprotein N gene (full length), the envelope protein E gene (full length), and the open reading frame1ab (ORF1ab) gene fragment (genomic position: 13201-15600, GenBank No.NC_045512). In the material, the copy number of ORF1ab gene was about 9.00◊ 10^5^ copies/µl, and the copy number of N gene was about 7.00×10^5^ copies/µl.

### 2.2 Primers and probes

Two genes of SARS-CoV-2 were used in rRT-PCR and ddPCR: an open reading frame1ab (ORF1ab) gene and nucleoprotein N gene. For targeting ORF1ab gene, we used the primers and probes used in previous research^[21]^. For targeting N gene, we used 5 sets of primers/probes, N(CDC) and N(USA) were referred from previous research (Laboratory testing for 2019-nCoV in humans) whereas N(LZU1), N(LZU2) and N(LZU3) were designed by ourselves. The sequences are described in Table 1. Primers and probes were purchased from The Beijing Genomics Institute (Beijing, China).

**Table1.**
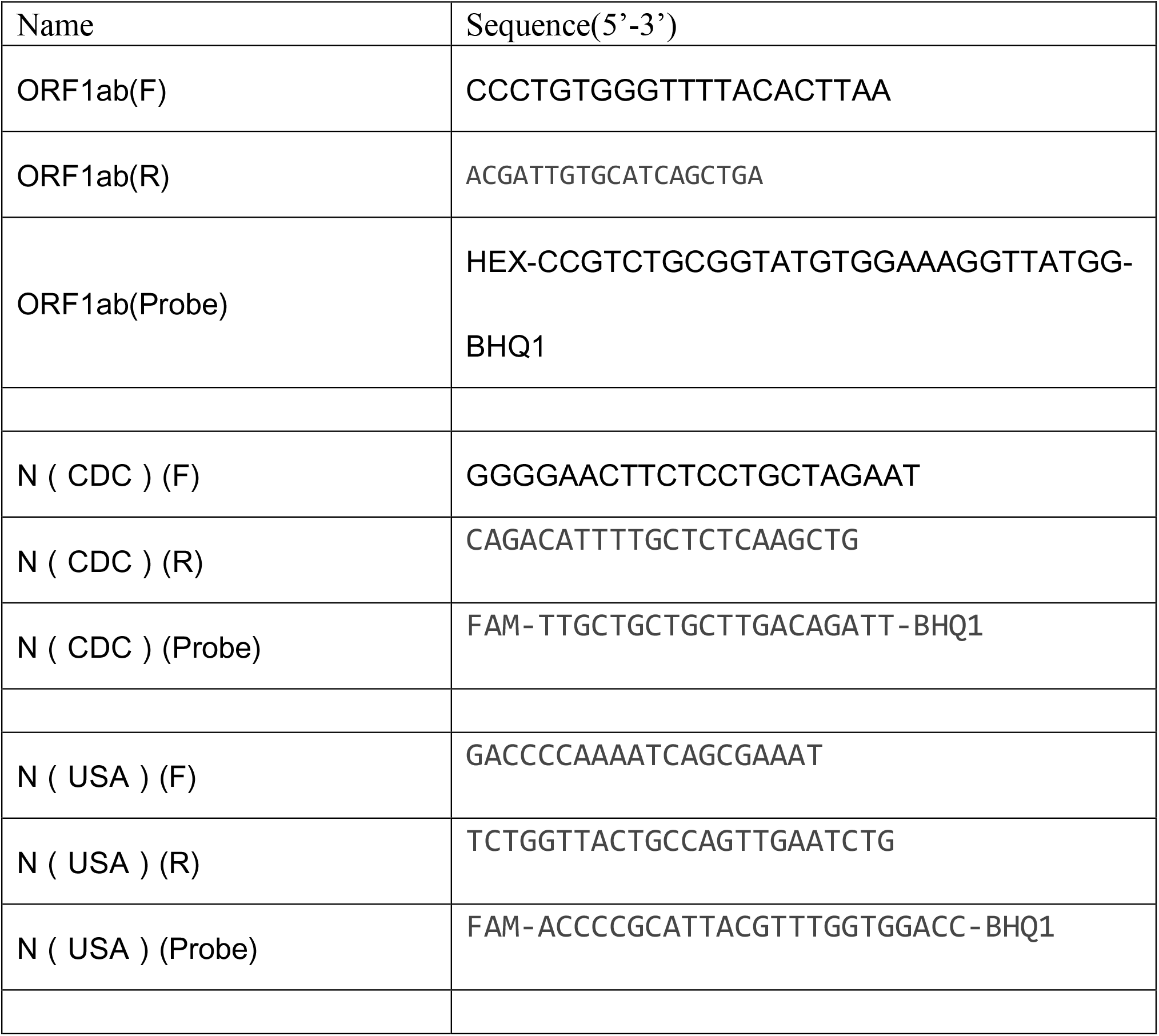
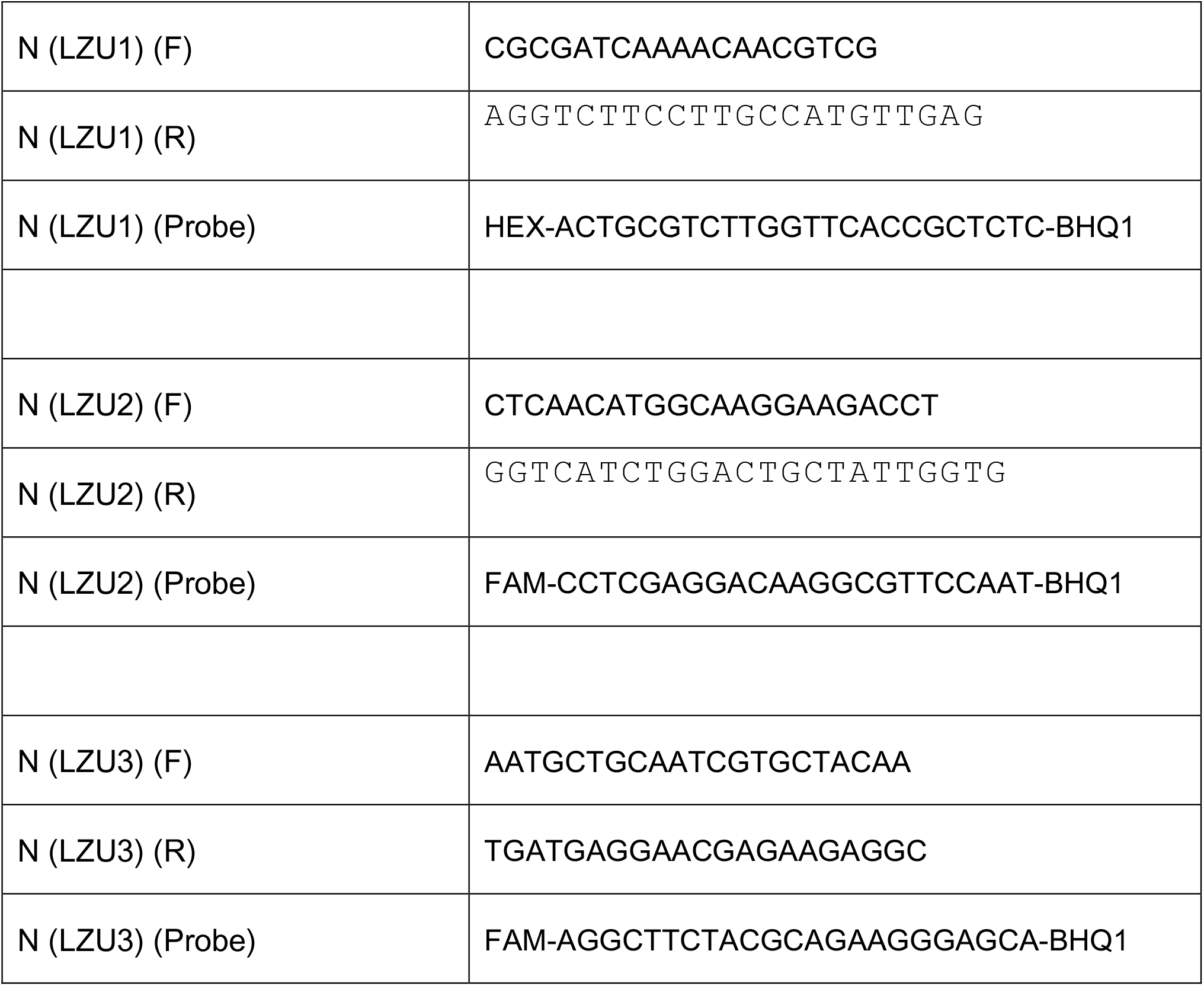
Primers and probes used in rRT-PCR and ddPCR

### 2.3 Multiplex rRT-PCR

An Applied Biosystems StepOnePlus™ Real-Time PCR Systems (Applied Biosystems, USA) was used for rRT-PCR. HI Script II U^+^ One Step rRT-PCR Probe Kit (Vazyme Biotech Co., Ltd, Nanjing, China) were used for rRT-PCR. Each 30 μL reaction mixture contained 15 μL of 2× One-Step U^+^ Mix, 1.5μL One-Step U^+^ Enzyme Mix, 0.6μL 50×ROX Reference Dye 1, 0.6μL Gene Specific Primer Forward (10μM),0.6μL of Gene Specific Primer Reverse (10μM), 0.3μL Taqman Probe (10μM), and 3 μL of total nucleic acid. The final concentrations of primers and probes were 0.2 μM and 0.1 μM, respectively. The standard cycling procedure was as follows: reverse transcription at 55°C for 15 min; polymerase activation at 95°C for 30 secs; and 45 cycles of PCR at 95°C for 10 secs and 60°C for 30 secs (signal acquisition). The filter combinations were 465-510 (FAM) and 540-580 (VIC). The Cq values were determined by the automated Fit Points method.

### 2.4 Droplet Digital PCR

For droplet digital PCR, we used the QX200 Droplet Digital PCR System and the One-Step RT-ddPCR Advanced Kit for Probes (Bio-Rad). First, we prepared the mixture of reaction, each 20 μL reaction mixture contained 5 μL 1× supermix, 2 μL Reverse Transcriptase (final concentration is 20U/μL), 1μL 300 mM DTT (final concentration is 15 mM), 1.8μL Gene Specific Primer Forward (10μM), 1.8μL of Gene Specific Primer Reverse (10μM), 0.5μL Taqman Probe (10μM), and 3 μL of total nucleic acid. The final concentrations of primers and probes were 900 nM and 250 nM, respectively. Secondly, twenty microliters of each reaction mix were converted into droplets with the QX200 droplet generator (Bio-Rad). Thirdly, the droplet-partitioned samples were then carefully transferred to a 96-well plate, sealed and cycled in a T100 Thermal Cycler (Bio-Rad) under the following cycling procedure: 50 °C for 60 min (Reverse Transcription), 95 °C for 10 min (Enzyme activation), followed by 40 cycles of 94 °C for 30 s (denaturation) and 55 °C for 1 min (annealing), then 98 °C for 10 min (enzyme deactivation), 4 °C for 30 min (droplet stabilization), and infinite 4-degree hold. Finally, after amplification, the cycled plate was then transferred and read in the FAM and HEX channels using the QX200 reader (Bio-Rad). The absolute copy numbers values were determined by the manual threshold line.

### 2.5 Data statistical analysis

Analysis of the ddPCR data was performed with Quanta Soft analysis software v.1.7.4.0917 (Bio-Rad) that accompanied the droplet reader to calculate the concentration of the target DNA sequences, along with their Poisson-based 95 % confidence intervals. The concentration reported by QuantaSoft equals to the copies of the template per microliter of the final 1× ddPCR reaction, which was also used in all the results. In addition, plots of linear regression were conducted with GraphPad Prism 7.00, and probit analysis for lower limit of detection (LLoD) was conducted with StatsDirect software v3.2.9. The LLoD were defined as the lowest concentration at which 95 % of positive samples were detected.

## 3. Results

### 3.1 One-step rRT-PCR reagents and probe chemistries

Simplex rRT-PCR with different set of primers/probe was performed using a synthetic standard RNA transcript with about 90000 copies/µl for ORF1ab and 70000 copies/µl for N gene as the rRT-PCR template. Each experiment was performed in duplicate. We compared the amplification efficiency and the average Ct values for target ORF1ab, N(CDC), N(USA1), N(LZU1), N(LZU2) and N(LZU3). The average Ct values and standard deviations (SD) were 23.293 (0.144), 24.113(1.059), 25.866(1.769), 23.682(0.399), 24.930(0.028), 25.919(0.039), respectively (data not shown). These results showed that the kit, the primers/probe set we designed, as well as the amplification condition, are workable.

### 3.2 Limit-of-detection (LOD) of simplex one-step RT-PCR with different set of primers/probes for target N gene

In order to investigate which primer/probe set has good amplification for the target gene, the LOD was analyzed using serially diluted synthetic SARS-CoV-2 RNA material (21000, 2100, 210, 105, 52.5, 21 and 2.1 copies/reaction for N gene). The Cq values, detection rates, slope, Y-intercept, R^2^, efficiency, and LOD with 95% detection probability are shown in **Table 2**. The standard curves for each primer/probe set were shown in **fig1**. Comparing with the N(CDC) and N(USA1), N(LZU3) has the similar Ct value and detection rate in the lower concentration of template, and the slope, Y-intercept, R^2^ of N(LZU3) were −3.228, 38.735, and 0.989, respectively (**fig 2**). The calculated PCR efficiency by machine was 104.093. The detection rate was 100% up to 105 copies/reaction. The calculated lower limit of detection (LOD) with 95% detection probability was 118 copies/reaction, which is the almost same as that of N (CDC) (121 copies/reaction) and N (USA1) (119 copies/reaction), suggesting that the prime/probe for N(LZU3) might be another potential primer/probe testing method for target gene N of SARS-CoV-2.

**Table 2.**
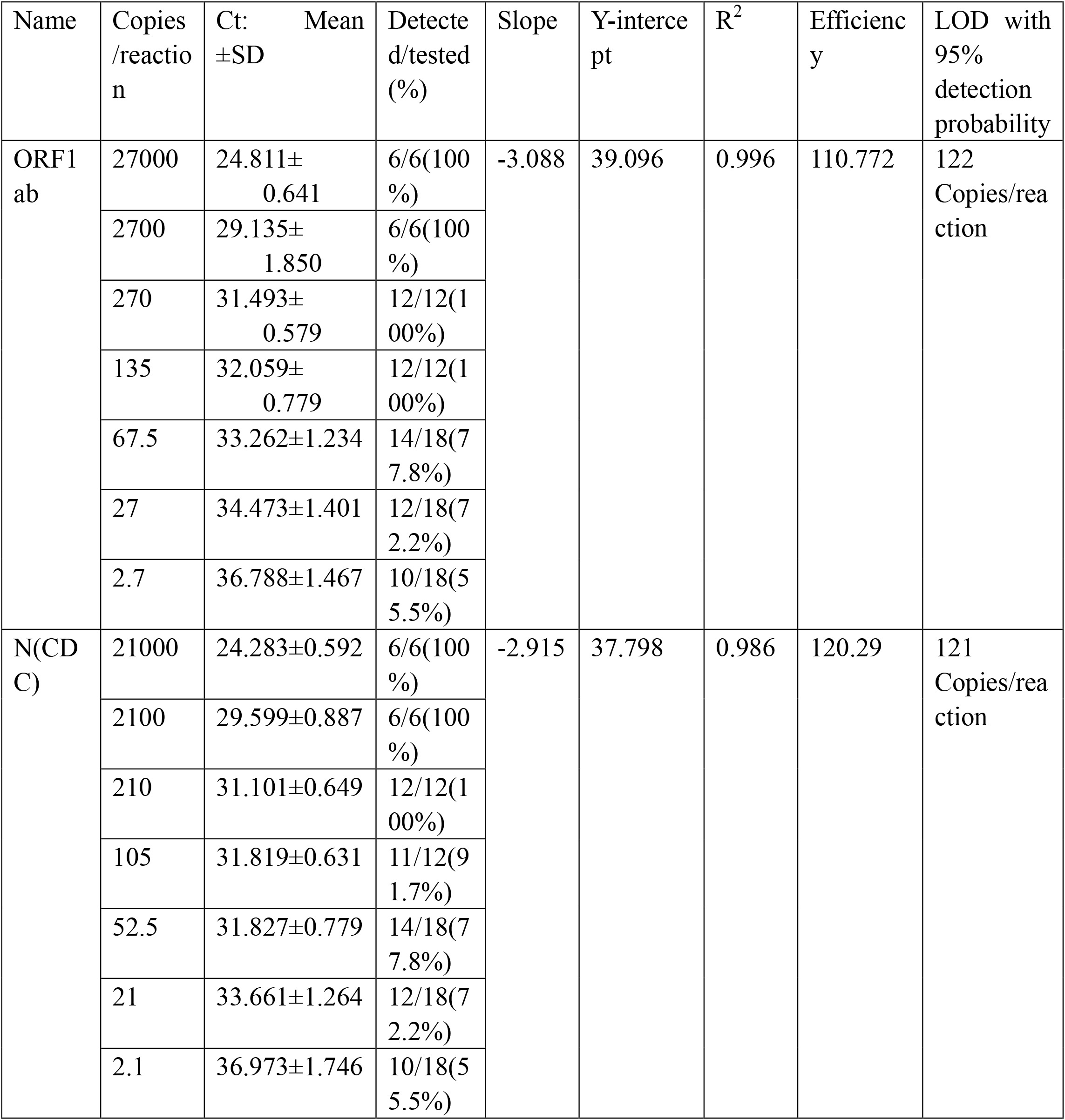
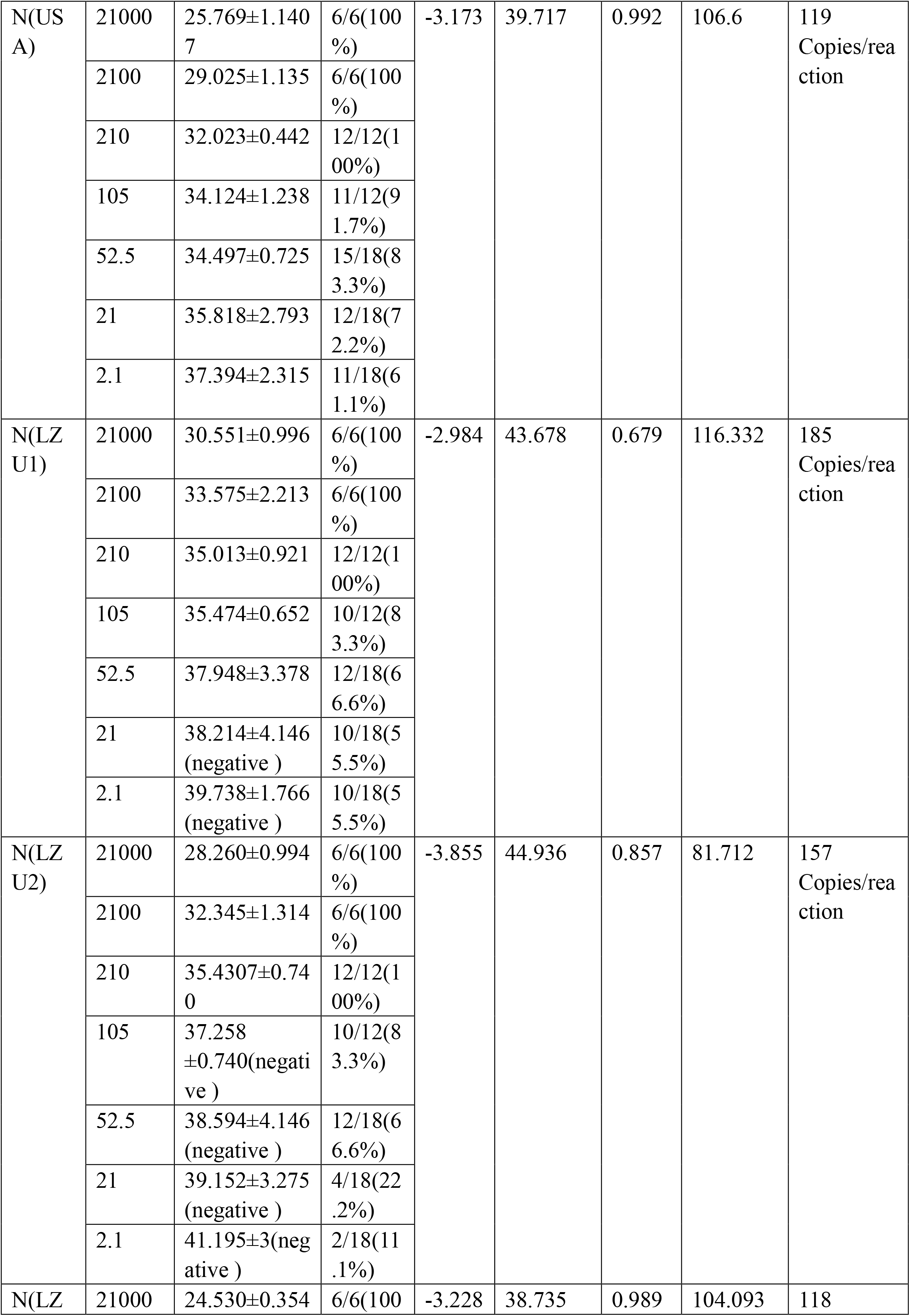
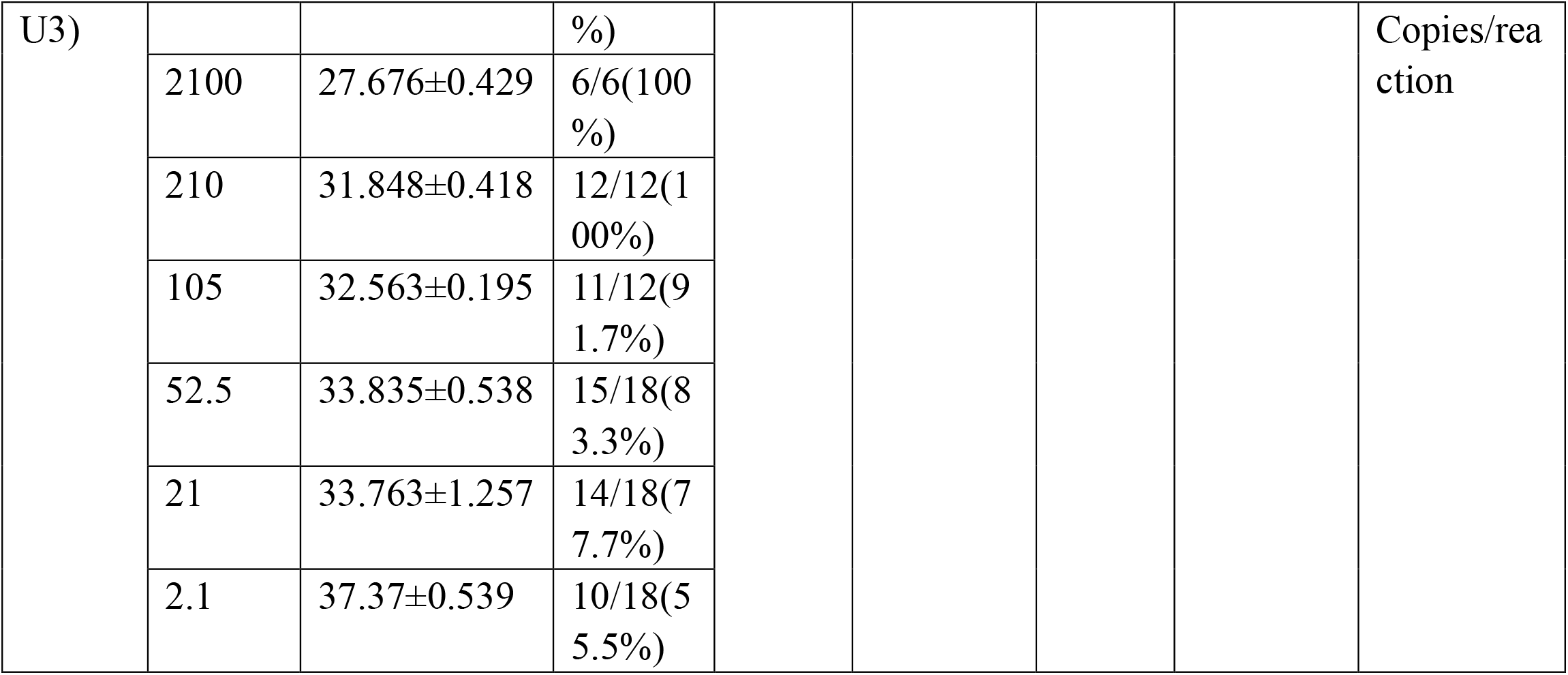
Sensitivity of simplex rRT-PCR with different primer and probe sets for N gene

**Figure 1.**
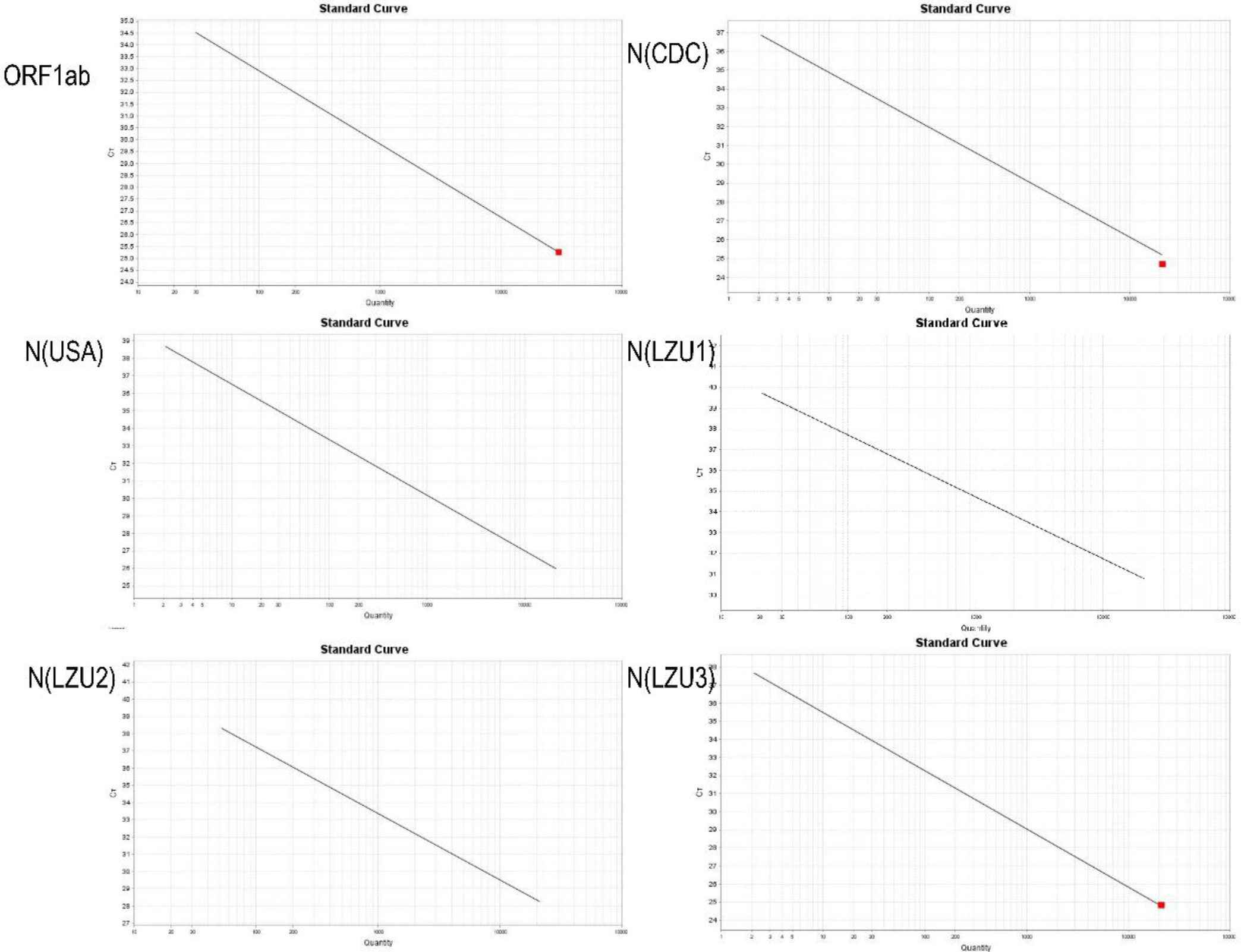
The standard curve of rRT-PCR using different primer/probe set target ORF1ab, N(CDC), N(USA), N(LZU1), N(LZU2), and N(LZU3). These maps were auto generated after amplification and download in the machine software. The mean Slope, Y-intercept, and R2 Efficiency for each primer/probe set were calculated in table 2. Data are representative of three independent experiments with 3 replicates for each copy.

**Figure2.**
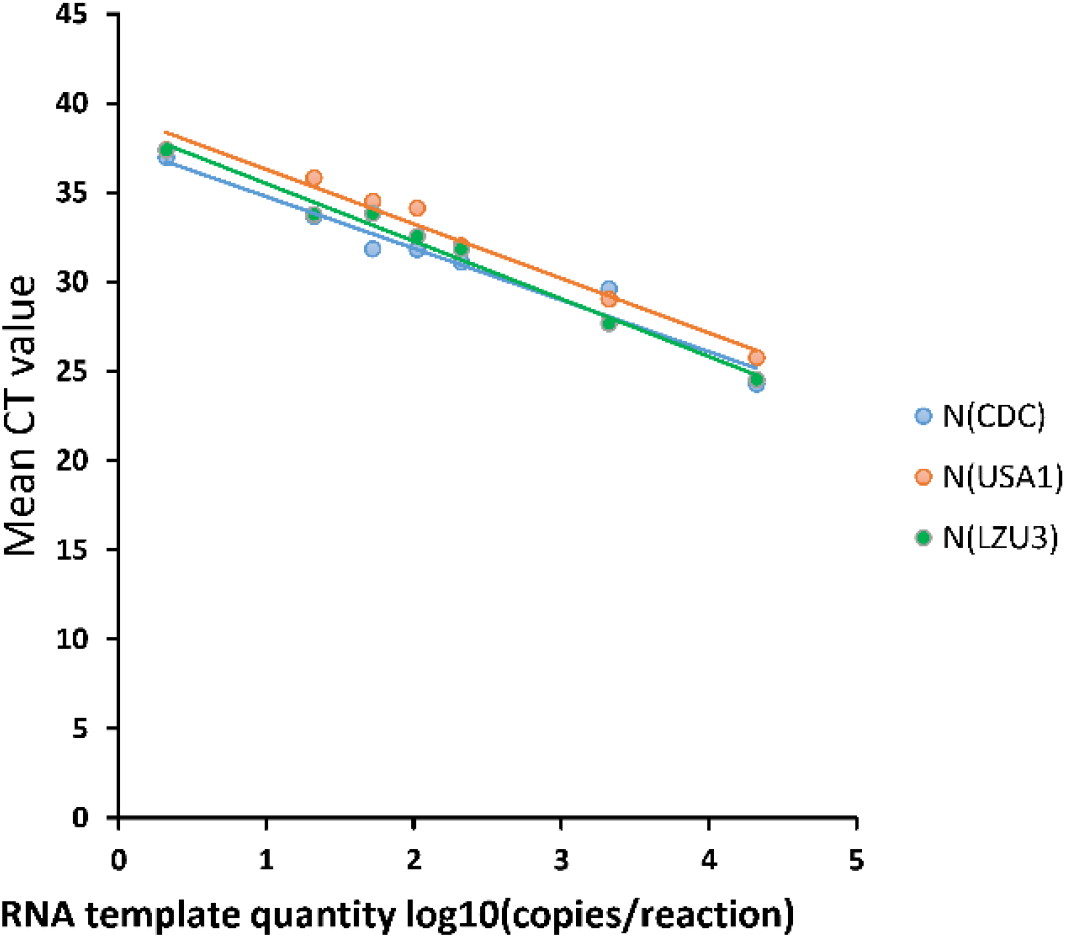
Plot of results from a linearity experiment to compare reportable range of rRT-PCR targeting for N of SARS-CoV-2 with different primer/probe set N(CDC), N(USA) and N(LZU3). RNA template quantity values (converted to log10) were plotted on the X axis versus measured Ct values (converted to log10) on the Y axis using EXCEL software. Data are representative of three independent experiments with 3 replicates for each concentration.

### 3.3 Detection of SARS-CoV-2 invitro transcribed RNA reference material with ddPCR

In order to investigate the detection efficiency of ddPCR on SARS-CoV-2, serial dilutions of a SARS-CoV-2 invitro transcribed RNA reference material (27000, 2700, 270, 135, 67.5, 27, 2.7 copies/reaction for ORF1ab gene and 21000, 2100, 210, 105, 52.5, 21, 2.1 copies/reaction for N gene) were tested with primers/probe sets ORF1ab, N (CDC), N(USA) and N(LZU3), the sequence can be seen in table 1. The ddPCR results were analyzed with Quanta software using a manual threshold to define the positive and negative droplets, the total events are above 15000 for each sample (data not shown), which make the statistics reasonable, and the positive and negative droplets could be distinctively separated (**fig 3**).

**Figure 3.**
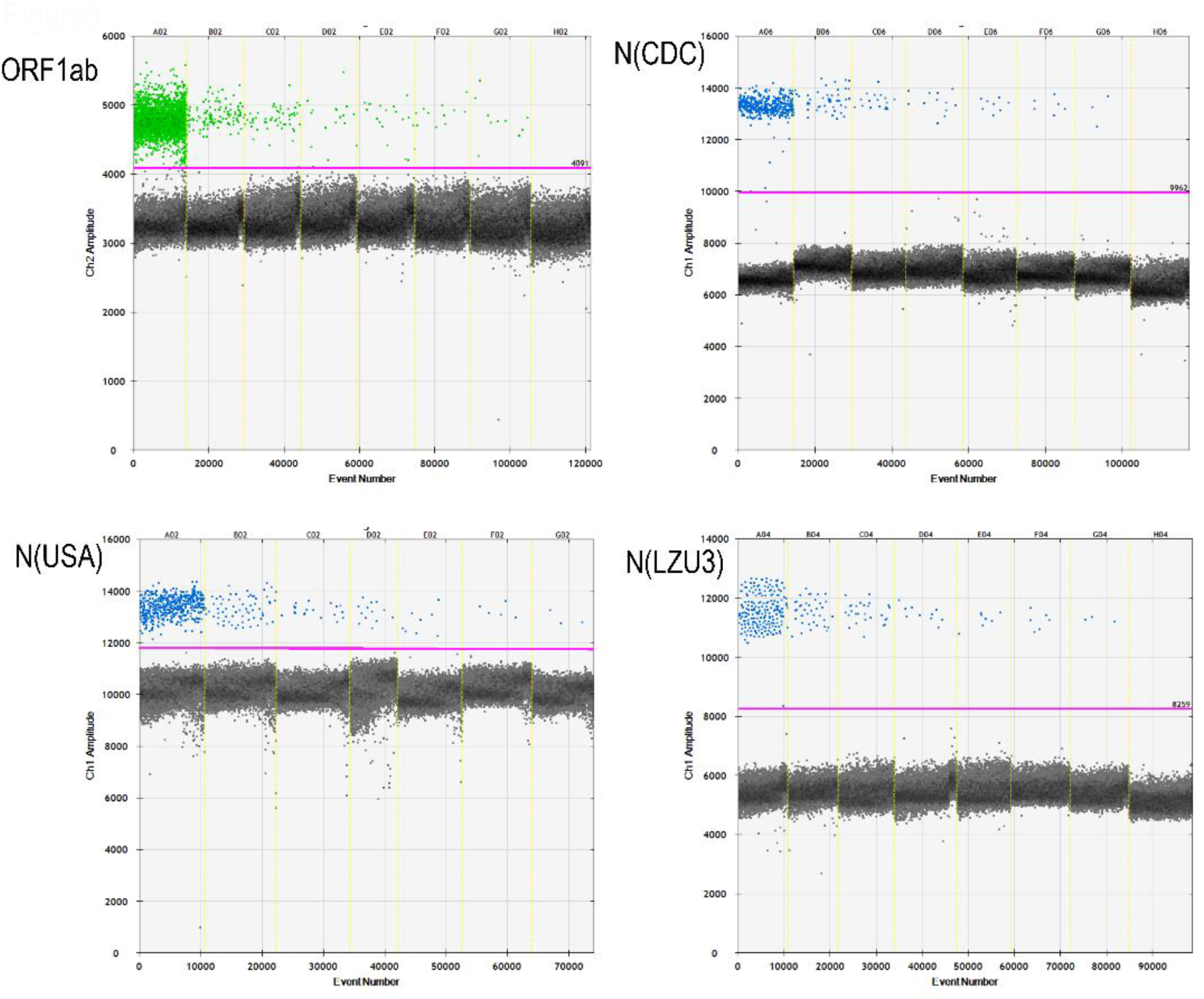
The ddPCR result (1D Amplitude) analyzed by QuantaSoft Version1.7.4.0917. Serial dilution of RNA template was detected with different primer/probe sets ORF1ab, N (CDC), N (USA), and N (LZU3). Column A to G means serial dilutions of 27000, 2700, 270, 135, 67.5, 27, and 2.7 copies/reaction for ORF1ab, and 21000, 2100, 210, 105, 52.5, 21and 2.1 copies/reaction for ORF1ab. Colunm H is represented the negative control (NTC, no template). The dark dots are represented the negative droplets, while the green and blue dots representative the positives droplets in different channels with probe HEX or FAM. The pink line is the manual multi well thresholding.

Linear regression was analyzed according to the mean measured quantity for each primer/probe sets. In **figure 4**, It shows good linearity for primer/probe sets ORF1ab, N(CDC), N(USA) and N(LZU3), (R2: 0.9917, 0.9886, 0.9978 and 0.9863, respectively). The reportable range of ddPCR is from 2.7copies/reaction to 27000 copies/reaction for ORF1ab and from 2.1copies/reaction to 21000 copies/reaction for N primes/probe sets. According to statistics, the Lower limit of detection (LOD) which was defined as the lowest concentration at which 95 % of positive samples were detected for primer/probe sets ORF1ab, N(CDC), N(USA) and N(LZU3) were 21.5 copies/reaction, 22.7 copies/reaction, 23.1 copies/reaction, and 22.4 copies/reaction, respectively, that are 1.075 copies/μL, 1.135 copies/μL, 1.155 copies/ μL, 1.12 copies/μL. In contrast, the LOD with 95% detection probability of rRT-PCR is from 118 copies/reaction to 121 copies/reaction for both ORF1ab and N primes/probe sets (table 2). Therefore, the ddPCR is more sensitive for samples with a low level quantity of samples.

**Figure 4.**
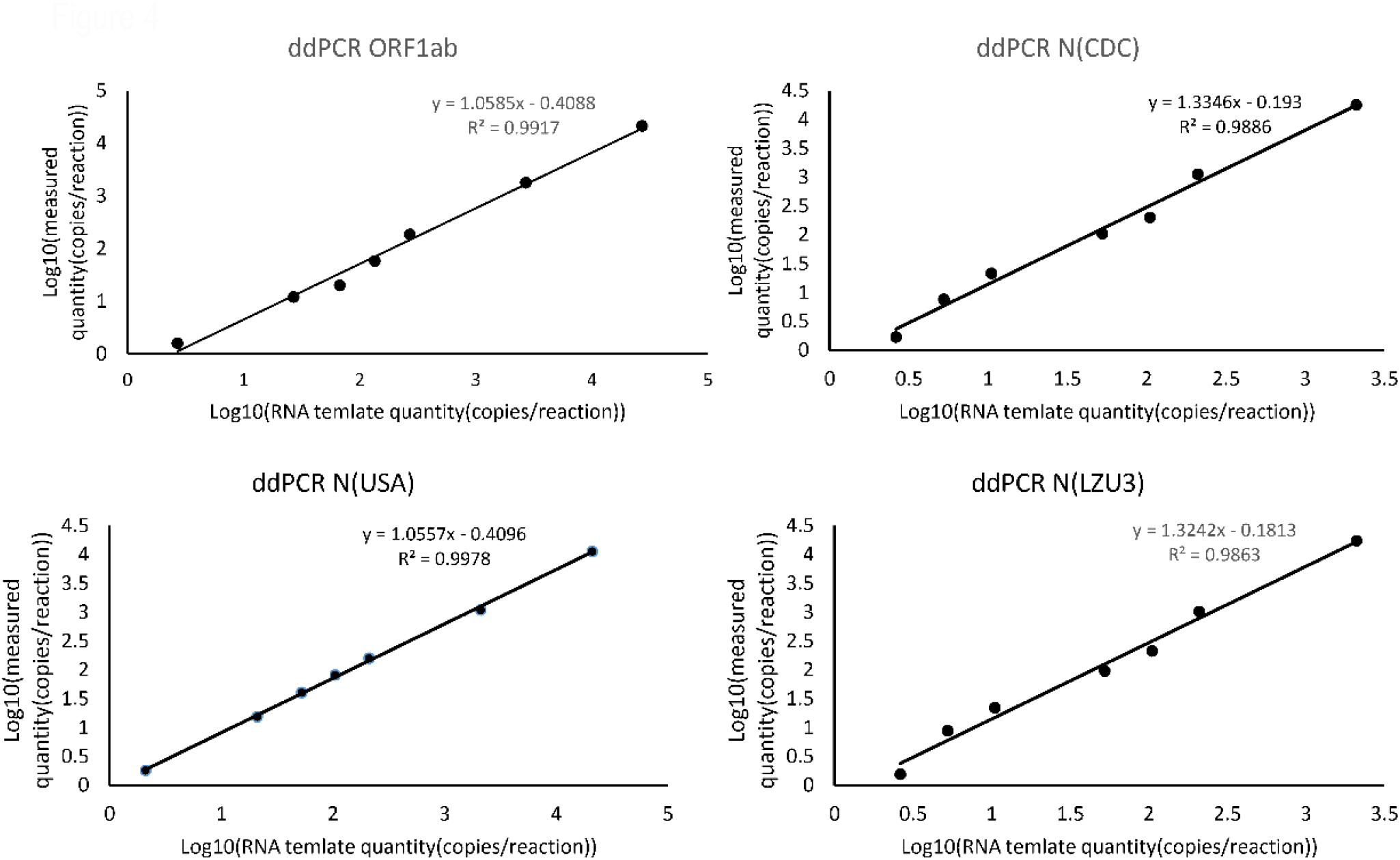
Plot of results from a linearity experiment to determine reportable range of ddPCR targeting for ORF1ab and N of SARS-CoV-2 using different primer/probe set. RNA template quantity values (converted to log10) were plotted on the X axis versus measured quantities (converted to log10) on the Y axis using EXCEL software. Data are representative of three independent experiments with 3 replicates for each concentration.

## Discussion

We developed a new primer/probe set N (LZU3) for targeting N gene of SARS-CoV-2 for molecular diagnosis of COVID-19 in both rRT-PCR and ddPCR. The sensitivity and specificity of rRT-PCR is affected by various factors, such as the nucleic acid extraction method, the one-step rRT-PCR reagent, and the primer/probe sets. Optimizing protocols are required before using this methodology in diagnostic laboratories ^[19]^. There are more asymptomatic infection and mild symptomatic infection in which the virus quantity might be very low, multiplex ddPCR might be much more useful for detecting asymptomatic infection.

We designed three primers/probes sets targeting the N gene of SARS-CoV-2. According to linear regression statistic in rRT-PCR, when the quantity of template was 105 copies/reaction, the detection rate for N(LZU1) and N(LZU2) were 83.3%, while the detection rate for N(LZU3) was 91.7%, which was similar with that of N(CDC) and N(USA) (91.7%) (table 2). Most importantly, if the quantity of template was 52.5 copies/reaction, the mean Ct value of N(LZU3) was 33.835, however, the mean Ct value of N(LZU1) and N(LZU2) were above 37, which would be assumed to be negative (table 2). For another hand, when we use N(LZU3) in ddPCR, it also showed good linearity with LOD at 22.4 copies/reaction. These result suggested that N (LZU3) might be another potential primer/probe set for detection of the SARS-CoV-2. Since the N (LZU3) target the different fragment N gene from N(CDC) and N(USA), these three primers/probe sets could be combine used in detection which may ensure multiplex rRT-PCR methodology highly sensitive for detection of SARS-CoV-2.

We also demonstrated that ddPCR had better sensitivity with lower LOD than rRT-PCR. We chose different primer/probe sets N (CDC), N (USA) and N (LZU3) for rRT-PCR, we found that the detection efficiency for low quantity of RNA template (lower than 52.5 copies/reaction) were limited (lower than 85%). The LOD with 95% detection probability for rRT-PCR was above 100 copies/reaction, which is 33.3 copies/ul. In this scenario we used one-step ddPCR to detect the same serial dilution of RNA template. The LOD with 95% detection probability for ddPCR was about 20 copies/reaction, which is about 1 copy/ul, which was 5 times lower than rRT-PCR These results confirmed that the ddPCR is more sensitive for low level quantity of sample which is similar to previously reported results. ^[22, 23]^. Moreover, the Biorad Company had investigated new kit (The Bio-Rad SARS-CoV-2 ddPCR Test) for detecting 2019-nCoV_N1, 2019-nCoV_N2, and Human RPP30 genes using Droplet Digital PCR. In the kit, the LoD for the samples extracted with the QIAamp Viral RNA Mini kit was also 625 cp/mL for N1 and N2, which were similar to our results.

In order to improve the detection sensitivity and specificity of SARS-CoV-2, at least two pairs of primer/probe sets were recommended for multiplex rRT-PCR or ddPCR. Our research demonstrated a pair of new primer/probe set for targeting N gene of SARS-CoV-2, which would be provide a complementary for multiplex detection of SARS-CoV-2.

Comparing with the suggested methods (rRT-qPCR. next generation sequencing (NGS), and immunological detection of IgM and IgG) for detection of SARS-CoV-2, ddPCR had several advantages. First of all, nucleic acid detecting is the most direct way to demonstrate the presence of the virus. Secondly, ddPCR may be more proper in which nucleic acid detection is the most direct way to demonstrate the presence of the virus ^[24]^, it could achieve absolute quantification of SARS-CoV-2 without the standard curve^[24]^. Thirdly, through detection within portioned droplets, ddPCR is much more sensitive ^[25]^ and accurate^[26]^ in portioned droplets detection, which is might be a strong tool for the detection of very low viral load in mild symptomatic and asymptomatic infection, it is also suitable for monitoring the change of the viral load in the convalescent patients^[27]^. Fifthly, ddPCR has a good tolerance for PCR inhibitory substances that might exist in the sample and reaction, therefore improve the detecting efficiency^[28]^. An additional advantage of ddPCR is that it can provide high concordance between sites, runs and operators^[26, 29, 30]^. However, it is not possible to compare Ct values on different runs or different machines. Thus, the established digital PCR method in this study with primer/probe N(LZU3) could be a powerful complementary method for detecting SARS-CoV-2.

The only limitation of the assay is the lack of clinical samples which required biosafety level 3 laboratory, however in future with the building of P3 facility, we will continue our work on ddPCR detection of SARS-CoV-2.

## Conclusion

We developed another potential primer/probe set targeting N gene for the detection of SARS-CoV-2, it could be used in both rRT-PCR and ddPCR. It can be combined with ORF1ab, N(CDC) or N(USA) in multiplex rRT-PCR and ddPCR. By using these primer/probe sets we have compared the analytical performance of rRT-PCR and ddPCR, we found that ddPCR could significantly improve the sensitivity of detection, which might be useful in detecting very low viral load in mild symptomatic and asymptomatic infection of COVID-19. Furthermore, it is also suitable for monitoring the change of the viral load in the convalescent patients.

## Data Availability

all data referred to in the manuscript are available.

## Reference

1. Valencia DN: Brief Review on COVID-19: The 2020 Pandemic Caused by SARS-CoV-2. Cureus 2020, 12(3):e7386.

2. Wang H, Li X, Li T, Zhang S, Wang L, Wu X, Liu J: The genetic sequence, origin, and diagnosis of SARS-CoV-2. European journal of clinical microbiology & infectious diseases : official publication of the European Society of Clinical Microbiology 2020.

3. Sohrabi C, Alsafi Z, O’Neill N, Khan M, Kerwan A, Al-Jabir A, Iosifidis C, Agha R: World Health Organization declares global emergency: A review of the 2019 novel coronavirus (COVID-19). International journal of surgery 2020, 76:71–76.

4. <20200816-covid-19-sitrep-209.pdf>.

5. Sharma R, Agarwal M, Gupta M, Somendra S, Saxena SK: Clinical Characteristics and Differential Clinical Diagnosis of Novel Coronavirus Disease 2019 (COVID-19). 2020:55–70.

6. Udugama B, Kadhiresan P, Kozlowski HN, Malekjahani A, Osborne M, Li VYC, Chen H, Mubareka S, Gubbay JB, Chan WCW: Diagnosing COVID-19: The Disease and Tools for Detection. 2020, 14(4):3822–3835.

7. Pfefferle S, Reucher S, Nörz D, Lütgehetmann M: Evaluation of a quantitative RT-PCR assay for the detection of the emerging coronavirus SARS-CoV-2 using a high throughput system. Euro surveillance : bulletin Europeen sur les maladies transmissibles = European communicable disease bulletin 2020, 25(9).

8. Hecht LS, Jurado-Jimenez A, Hess M, Halas HE, Bochenek G, Mohammed H, Alzahrani F, Asiri MO, Hasan R, Alamri A et al: Verification and diagnostic evaluation of the RealStar((R)) Middle East respiratory syndrome coronavirus (N gene) reverse transcription-PCR kit 1.0. Future microbiology 2019, 14:941–948.

9. Hong TC, Mai QL, Cuong DV, Parida M, Minekawa H, Notomi T, Hasebe F, Morita K: Development and evaluation of a novel loop-mediated isothermal amplification method for rapid detection of severe acute respiratory syndrome coronavirus. Journal of clinical microbiology 2004, 42(5):1956–1961.

10. Balboni A, Gallina L, Palladini A, Prosperi S, Battilani M: A real-time PCR assay for bat SARS-like coronavirus detection and its application to Italian greater horseshoe bat faecal sample surveys. TheScientificWorldJournal 2012, 2012:989514.

11. Schwartz SL, Lowen AC: Droplet digital PCR: A novel method for detection of influenza virus defective interfering particles. Journal of virological methods 2016, 237:159–165.

12. Hu W, Bai B, Hu Z, Chen Z, An X, Tang L, Yang J, Wang H, Wang H: Development and evaluation of a multitarget real-time Taqman reverse transcription-PCR assay for detection of the severe acute respiratory syndrome-associated coronavirus and surveillance for an apparently related coronavirus found in masked palm civets. Journal of clinical microbiology 2005, 43(5):2041–2046.

13. LeBlanc JJ, Gubbay JB, Li Y, Needle R, Arneson SR, Marcino D, Charest H, Desnoyers G, Dust K, Fattouh R et al: Real-time PCR-based SARS-CoV-2 detection in Canadian laboratories. Journal of clinical virology : the official publication of the Pan American Society for Clinical Virology 2020, 128:104433.

14. Chan JF, Yip CC, To KK: Improved Molecular Diagnosis of COVID-19 by the Novel, Highly Sensitive and Specific COVID-19-RdRp/Hel Real-Time Reverse Transcription-PCR Assay Validated In Vitro and with Clinical Specimens. 2020, 58(5).

15. Wu F, Zhao S, Yu B, Chen YM, Wang W, Song ZG, Hu Y: A new coronavirus associated with human respiratory disease in China. Nature 2020, 579(7798):265–269.

16. Nalla AK, Casto AM, Huang MW, Perchetti GA, Sampoleo R, Shrestha L, Wei Y, Zhu H, Jerome KR, Greninger AL: Comparative Performance of SARS-CoV-2 Detection Assays Using Seven Different Primer-Probe Sets and One Assay Kit. Journal of clinical microbiology 2020, 58(6).

17. Bordi L, Piralla A, Lalle E, Giardina F, Colavita F, Tallarita M, Sberna G, Novazzi F, Meschi S, Castilletti C et al: Rapid and sensitive detection of SARS-CoV-2 RNA using the Simplexa™ COVID-19 direct assay. Journal of clinical virology : the official publication of the Pan American Society for Clinical Virology 2020, 128:104416.

18. Choudhary ML, Vipat V, Jadhav S, Basu A, Cherian S, Abraham P, Potdar VA: Development of in vitro transcribed RNA as positive control for laboratory diagnosis of SARS-CoV-2 in India. The Indian journal of medical research 2020, 151(2 & 3):251-254.

19. Ishige T, Murata S, Taniguchi T, Miyabe A, Kitamura K, Kawasaki K, Nishimura M, Igari H, Matsushita K: Highly sensitive detection of SARS-CoV-2 RNA by multiplex rRT-PCR for molecular diagnosis of COVID-19 by clinical laboratories. Clinica chimica acta; international journal of clinical chemistry 2020, 507:139–142.

20. Wang X, Yao H, Xu X, Zhang P, Zhang M, Shao J, Xiao Y, Wang H: Limits of Detection of Six Approved RT-PCR Kits for the Novel SARS-coronavirus-2 (SARS-CoV-2). Clinical chemistry 2020.

21. Corman VM, Landt O, Kaiser M, Molenkamp R, Meijer A, Chu DK, Bleicker T, Brunink S, Schneider J, Schmidt ML et al: Detection of 2019 novel coronavirus (2019-nCoV) by real-time RT-PCR. Euro surveillance : bulletin Europeen sur les maladies transmissibles = European communicable disease bulletin 2020, 25(3).

22. Tao Suo XL, Ming Guo, Jiangpeng Feng,Wenjia Hu, Yang Yang, Qiuhan Zhang, Xin Wang, Muhanmmad Sajid,Dong Guo,Zhixiang Huang,Liping Deng,Tielong Chen,Fang Liu,Xu Ke,Yuan Liu, Qi Zhang,Yingle Liu,Yong Xiong,Guozhong Chen, Yu Chen, Ke Lan: ddPCR: a more sensitive and accurate tool for SARS-CoV-2 detection in low viral load specimens. MedRxIv 2020.

23. Lianhua Dong JZ, Chunyan Niu, Quanyi Wang,Yang Pan,Xia Wang,Yongzhuo Zhang, Jiayi Yang, Manqing Liu, Yang Zhao,Tao Peng,Jie Xie,Yunhua Gao, Di Wang, Yun Zhao, Xinhua Dai,Xiang Fang: Highly accurate and sensitive diagnostic detection of SARS-CoV-2 by digital PCR. MedRxIv PRE-PRINT 2020.

24. Zhou D, Li Y, Li J, Yu J, Yang H, Wei H: Construction of Lentivirus-Based Reference Material for RT-PCR Detection of Middle East Respiratory Syndrome Coronavirus and Its Application in External Quality Assessment. Journal of nanoscience and nanotechnology 2019, 19(9):5510–5516.

25. Persaud D, Gay H, Ziemniak C, Chen YH, Piatak M, Jr., Chun TW, Strain M, Richman D, Luzuriaga K: Absence of detectable HIV-1 viremia after treatment cessation in an infant. The New England journal of medicine 2013, 369(19):1828–1835.

26. Whale AS, Devonshire AS, Karlin-Neumann G, Regan J, Javier L, Cowen S, Fernandez-Gonzalez A, Jones GM, Redshaw N, Beck J et al: International Interlaboratory Digital PCR Study Demonstrating High Reproducibility for the Measurement of a Rare Sequence Variant. Analytical chemistry 2017, 89(3):1724–1733.

27. Patterson B, Morrow C, Singh V, Moosa A, Gqada M, Woodward J, Mizrahi V, Bryden W, Call C, Patel S et al: Detection of Mycobacterium tuberculosis bacilli in bio-aerosols from untreated TB patients. Gates open research 2017, 1:11.

28. Dingle TC, Sedlak RH, Cook L, Jerome KR: Tolerance of droplet-digital PCR vs real-time quantitative PCR to inhibitory substances. Clinical chemistry 2013, 59(11):1670–1672.

29. Dong L, Wang X, Wang S, Du M, Niu C, Yang J, Li L, Zhang G, Fu B, Gao Y et al: Interlaboratory assessment of droplet digital PCR for quantification of BRAF V600E mutation using a novel DNA reference material. Talanta 2020, 207:120293.

30. Whale AS, Jones GM, Pavsic J, Dreo T, Redshaw N, Akyurek S, Akgoz M, Divieto C, Sassi MP, He HJ et al: Assessment of Digital PCR as a Primary Reference Measurement Procedure to Support Advances in Precision Medicine. Clinical chemistry 2018, 64(9):1296–1307.

